# Multi-omics analysis of serial samples from metastatic TNBC patients on PARP inhibitor monotherapy provide insight into rational PARP inhibitor therapy combinations

**DOI:** 10.1101/2020.07.25.20146431

**Authors:** Marilyne Labrie, Allen Li, Allison Creason, Courtney Betts, Jamie Keck, Brett Johnson, Shamilene Sivagnanam, Christopher Boniface, Hongli Ma, Aurora Blucher, Young Hwan Chang, Koei Chin, Jacqueline Vuky, Alexander R. Guimaraes, Molly Downey, Jeong Youn Lim, Lina Gao, Kiara Siex, Swapnil Parmar, Annette Kolodzie, Paul T Spellman, Jeremy Goecks, Lisa M. Coussens, Christopher L. Corless, Raymond Bergan, Joe W. Gray, Gordon B. Mills, Zahi I. Mitri

**Author notes:** **Corresponding authors**: Zahi I. Mitri, MD, MS, Oregon Health & Science University, 3181 SW Sam Jackson Park Rd., Mail Code: OC14HO, Portland, OR 97239, Marilyne Labrie, PhD, Oregon Health & Science University, 2720 S Moody Avenue, Mailcode: KR-PM, Portland, OR 97201.

## Abstract

Due to complexity of advanced epithelial cancers, it is necessary to implement patient specific combination therapies if we are to markedly improve patient outcomes. However, our ability to select and implement patient specific combination therapies based on dynamic molecular changes in the tumor and tumor ecosystem in response to therapy remains extremely limited. In a pilot study, we evaluated the feasibility of real-time deep analysis of serial tumor samples from triple negative breast cancer patients to identify mechanisms of resistance and treatment opportunities as they emerge under therapeutic stress engendered by poly-ADP-ribose polymerase (PARP) inhibitors (PARPi). Although PARP inhibition was consistently observed in all patients, deep molecular analysis of the tumor and its ecosystem revealed insights into potentially effective therapeutic PARPi combinations. In a BRCA-mutant basal breast cancer exceptional long-term survivor, we noted striking PARPi-induced tumor destruction accompanied by a marked infiltration of immune cells containing CD8 effector cells, consistent with pre-clinical evidence for association between STING mediated immune activation and benefit from PARPi and immunotherapy. Tumor cells in the exceptional responder underwent extensive protein network rewiring in response to PARP inhibition. In contrast, there were minimal changes in the ecosystem of a luminal androgen receptor (LAR) rapid progressor in response to PARPi likely due to indifference to the effects of PARP inhibition. In this rapid progressor, there was minimal evidence of immune activation or protein network rewiring in response to PARPi, despite PARP being inhibited, and no clinical benefit was noted for this participant. Together, deep real-time analysis of longitudinal biopsies identified a suite of PARPi-induced emergent changes including immune activation, DNA damage checkpoint activation, apoptosis and signaling pathways including RTK, PI3K-AKT and RAS-MAPK, that could be used to select patient specific combination therapies, based on tumor and immune state changes that are likely to benefit specific patients.

**Highlights:**

- Longitudinal analysis of serial tumor samples in real-time identifies adaptive mechanisms of resistance to PARPi therapies.
- Deep molecular analysis of the tumor reveals insights into potentially effective therapeutic PARPi combinations.
- Extensive protein network rewiring, microenvironment and immune state changes are assessable factors for patient specific combination therapies.

## Introduction

Triple negative breast cancer (TNBC) is characterized by lack of expression of estrogen (ER) and progesterone (PR) receptors and human epidermal growth factor receptor 2 (HER2). Despite advances in our understanding of TNBC biology, metastatic TNBC (mTNBC) remains an incurable disease, with limited therapeutic options and high mortality. A key to developing therapies that induce durable responses is the recognition that TNBC represents a molecularly heterogenous disease with several clinically relevant subtypes. Indeed, some of the “failures” of clinical trials in mTNBC may be due to the heterogeneous nature of the disease, such that therapies are not tested against the appropriate subsets. A precision oncology approach of targeting therapeutic liabilities as they arise in response to therapy in mTNBC may increase the efficiency of selection of patient specific combination therapy.

Recent clinical trials have aimed at capitalizing on molecular alterations in mTNBC to implement effective novel therapies. For patients with germline *BRCA* mutations, the PARP inhibitors olaparib and talazoparib are now FDA-approved, based on improved efficacy and safety of PARPi therapies compared to cytotoxic chemotherapies (Robson et al., 2017, Turner et al., 2019, Robson et al., 2019). Additionally, immune checkpoint blockade (ICB) therapy in combination with chemotherapy was approved for frontline treatment of mTNBC expressing PD-L1 based on improved progression-free and overall survival (Schmid et al., 2018, Schmid et al., 2020). However, despite these promising advances, virtually all patients develop progressive disease on PARPi- or ICB-based therapies, indicating a critical need to implement patient specific combinations that can interdict or overcome resistance as it emerges in mTNBC.

The combination of PARPi and ICB is effective in breast cancer models wild-type for *BRCA1/2* and competent for DNA damage repair. These models have shown induction of a stimulator of interferon genes (STING) response that induces interferon production and immune activation (Jiao et al., 2017, Chabanon et al., 2019, Ding et al., 2018, Shen et al., 2019). The MEDIOLA and TOPACIO trials both demonstrated that PARPi and ICB combinations are well tolerated and have clinical activity in a subset of both *BRCA-*mutant as well as *BRCA-*wild type solid tumors, albeit with much more limited activity in *BRCA*-wild type tumors (Konstantinopoulos, 2017, Domchek et al., 2019, Domchek et al., 2018). Unfortunately, serial biopsies were only available on a limited number of patients in these studies, precluding identification of predictive biomarkers and resistance mechanisms that enabled tumors to eventually escape control. This is especially important because biomarkers able to accurately predict benefit from PARPi mono or combination therapy in biopsies taken prior to treatment have not been identified.

Tumors and the tumor ecosystem rapidly adapt to therapeutic stress on treatment with therapeutic agents, including PARPi. These adaptive responses indicate both mechanisms of resistance and therapeutic opportunities. We and others have demonstrated that combination therapy designed to capitalize on adaptive responses induced by therapy can generate tumor control in preclinical model systems (Choi et al., 2016, Corcoran et al., 2018, Fang et al., 2018, Iavarone et al., 2019, Kurimchak et al., 2016, Labrie et al., 2019b, Zawistowski et al., 2017, Fang et al., 2019, Sun et al., 2017, Konstantinopoulos et al., 2019, Matulonis et al., 2017). We have translated many of these observations to clinical trials, with marked patient benefit (NCT03162627, NCT02208375, NCT02659241, NCT03586661, NCT02316834, NCT03544125, NCT03801369, and NCT03637491). While analysis of baseline and on-therapy biopsies provides significant information, the information content on changes induced by therapy appears to provide the most information for selection of combination therapies (Labrie et al., 2019b).

This pilot clinical trial evaluated the safety and efficacy of the combination of olaparib and durvalumab for treatment of mTNBC patients and feasibility of longitudinal analysis of serial tumor samples in real-time to identify adaptive mechanisms of resistance as they emerge under PARPi therapeutic stress. This study incorporated comprehensive multi-omics analysis of serial liquid and tumor biopsies under the precision oncology platform: Serial Measurements of Molecular and Architectural Responses to Therapy (SMMART) at the Oregon Health & Science University Knight Cancer Institute (OHSU KCI) (Mitri et al., 2018). The SMMART analytic platform and robust computational biology pipeline enables study of adaptive mechanisms of resistance as they emerge and offers a new design of biomarker-driven clinical trials aimed at identifying markers of benefit as well as selecting combinations of drugs in real-time to benefit the patient under therapy.

## Methods

### Study Design and Participants

This was a single-center, open-label, single-arm, pilot study of the PARPi olaparib and the PD-L1 antibody durvalumab **(Fig. 1A)**. Eligibility criteria included biopsy proven mTNBC, ECOG PS ≤2, received ≤2 chemotherapeutic courses in the metastatic setting. Patients with prior PARPi or ICB exposure in the metastatic setting were excluded. The study schema is summarized in Fig. 1A. The study was reviewed and approved by the OHSU Institutional Review Board.

**Figure 1:**
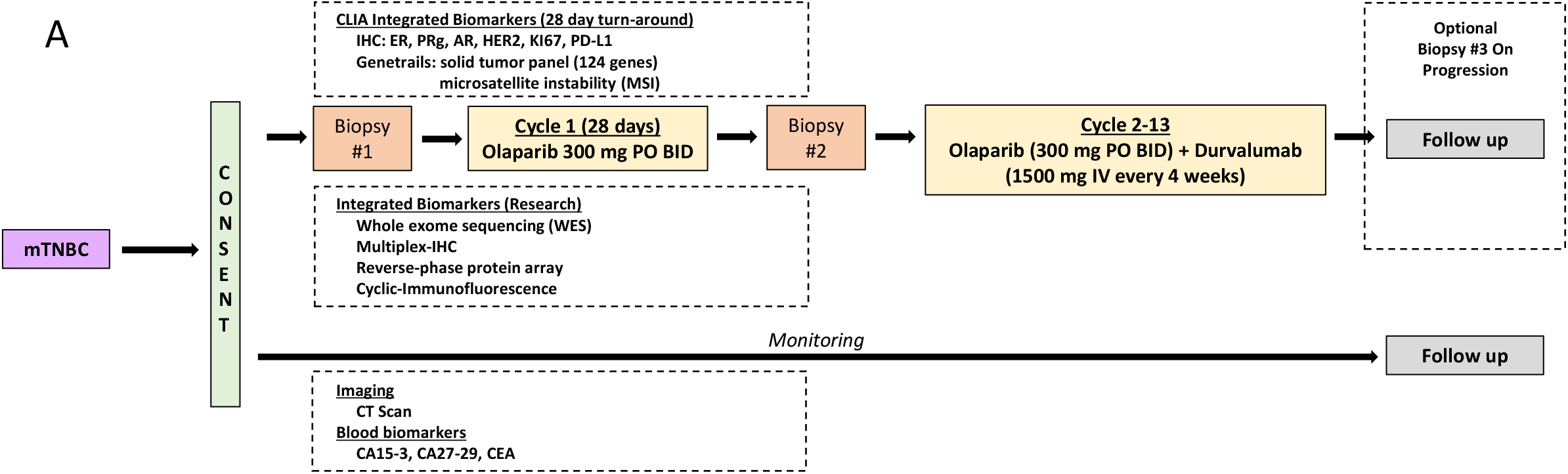
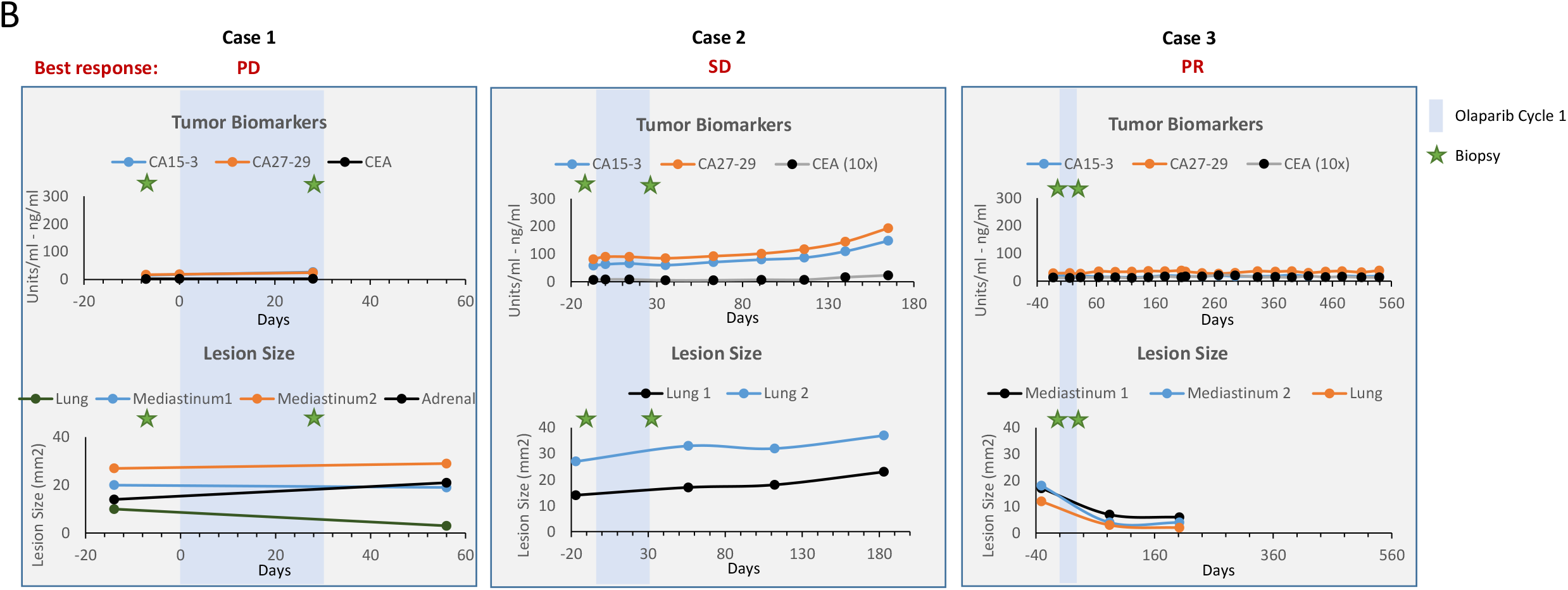
Schematic of study design and patient’s response to olaparib and durvalumab combination. **(A)** After consent, mTNBC patients were subjected to a pre-treatment biopsy (#1) followed by one cycle of olaparib (28 days) and an on-treatment biopsy (#2). Patents were then treated with a combination of olaparib and durvalumab for up to 12 additional cycles. **(B) (Upper panel)** Blood biomarkers, **(lower)** lesion size. Green stars represent the time of pre and on-treatment biopsies collection and the blue bar represent the first cycle of olaparib monotherapy, which started on day 0.

### Study Objectives

The primary objective was to assess the feasibility of completing a suite of Clinical Laboratory Improvement Amendments (CLIA) assays (IHC, Genetrail© Comprehensive Tumor Panel and serum biomarkers) and accompanying analytics (mIHC, RPPA, Cyc-IF) on pre-treatment biopsies within a planned 4-week window for enrolled participants. Secondary objectives included safety and tolerability of the combination; as well as preliminary efficacy endpoints (response rate per RECIST v1.1 and irRECIST, time to disease progression, and overall survival) of olaparib and durvalumab. Exploratory objectives included examining response rates depending on tumor characteristics, identifying candidate predictive biomarkers of sensitivity, identifying emerging candidate mechanisms of resistance to therapy, determining changes in tumor cells induced by PARPi, and identifying any candidate tumor markers that may suggest candidate combinatorial therapies for overcoming treatment resistance.

### Tumor Biopsies

Biopsies were performed under ultrasound or CT-guidance using 18 gauge needles. Between 3 and 6 passes were performed on a selected lesion and immediately handed off for: a) fixation in 10% neutral-buffered formalin; b) flash-freezing in liquid nitrogen; and c) fixation in glutaraldehyde. Formalin fixation was for 24 hours, after which cores were transferred to 70% ethanol and submitted for routine processing, paraffin embedding and sectioning in a CLIA histology laboratory.

### Immunohistochemistry (IHC)

Five micron sections of tumor biopsies were subjected to immunohistochemistry using Ventana reagents and a Ventana autostainer in a CLIA histology laboratory. Each biopsy, dependent upon sufficient sample, was assessed for the expression of ER, PR, AR, HER2 and Ki67.All stained sections, including H&Es, were scanned on a Leica AT2 digital scanner.

### Serum tumor markers

CA15-5, CA27-29 and CEA were measured using standard methodologies in a CLIA laboratory.

### GeneTrails© Comprehensive Solid Tumor Panel

This next-generation sequencing panel was run in the Knight Diagnostic Laboratories (CLIA-licensed/CAP-accredited). Tumor-rich areas of unstained five micron sections of FFPE biopsies were macro-dissected and nucleic acid was extracted using a commercial kit. Two separate amplicon-based libraries (one DNA, one RNA/cDNA) were sequenced and analyzed as detailed in **Suppl. method**. The DNA library covered 124 cancer-related genes and is used to detect single nucleotide variants, insertions/deletions, copy number alterations, and microsatellite instability.

### Whole exome DNA sequencing and somatic variant calling

Total DNA was isolated from FFPE biopsies using QIAgen FFPE DNA extraction kits. Whole-exome DNA sequencing libraries were prepared from 100-500 ng DNA using KAPA Hyper-Prep Kit (KAPA Biosystems) with Agilent SureSelect XT Target Enrichment System and Human All Exon V5 capture baits (Agilent Technologies). Next generation sequencing was carried out using the Illumina NovaSeq 6000 by Novogene, to produce 150bp paired end reads with an average depth of 500X tumor/cfDNA or 100X matched normal (buffy coat DNA). FastQ data files were aligned and processed using BWA MEM (0.7.12, GATK, Broad Institute). Somatic variants were called following the GATK Best Practices Somatic Short Variant Discovery, including GATK4 MuTect2 (2.1-beta, Broad Institute). Variants were selected based on the following criteria: tumor > 30X; normal site depth > 14X; tumor variant allele frequency > 5%; normal vaf < 2%; and sites found to be in dbSNP (https://www.ncbi.nlm.nih.gov/projects/SNP/) or a panel of normals were removed. All variants were hand-curated with Samtools mpileup (1.2, https://github.com/samtools/samtools) and Interactive Genome Viewer (2.3.82, Broad Institute) to assess their validity in context. Whole exome tumor copy numbers were estimated using CNVkit (0.9.4a0) of the tumor sample compared to a pool of blood normal samples.

### Multiplex Immunohistochemistry, Image Acquisition and Processing

Multiplex IHC was performed on 5 μm FFPE sections using an adapted protocol based on methodology described previously (Tsujikawa et al., 2017, Banik et al., 2020) and detailed in **Suppl. Method**. Primary antibody details, dilution, and incubation times are listed in **Suppl. Table 1**. Cells were classified based on hierarchical gating (image cytometry) and defined as described in **Fig. 2**. For visualization, signal-extracted images were overlaid in pseudo-color in FIJI.

**Figure 2:**
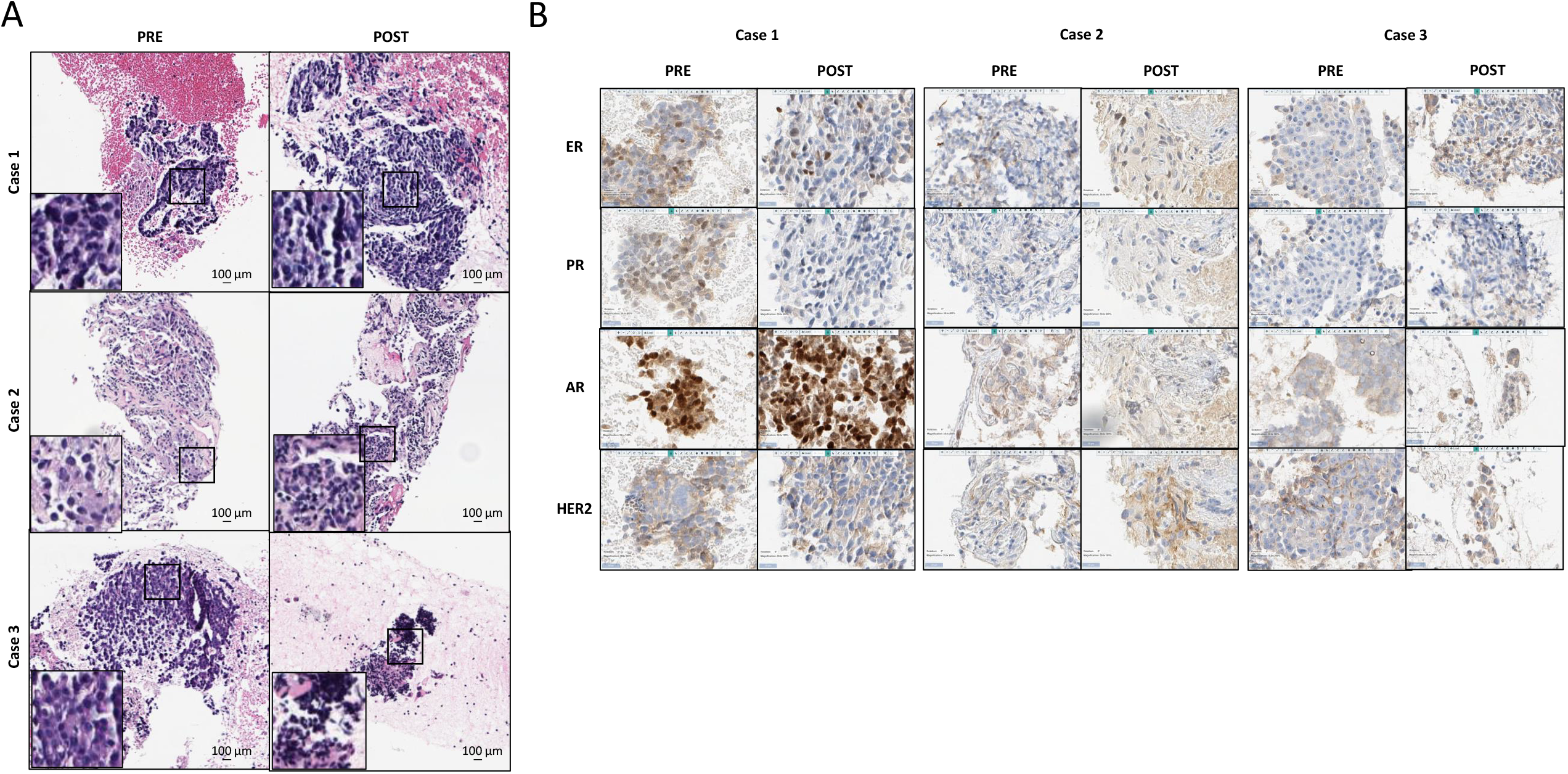
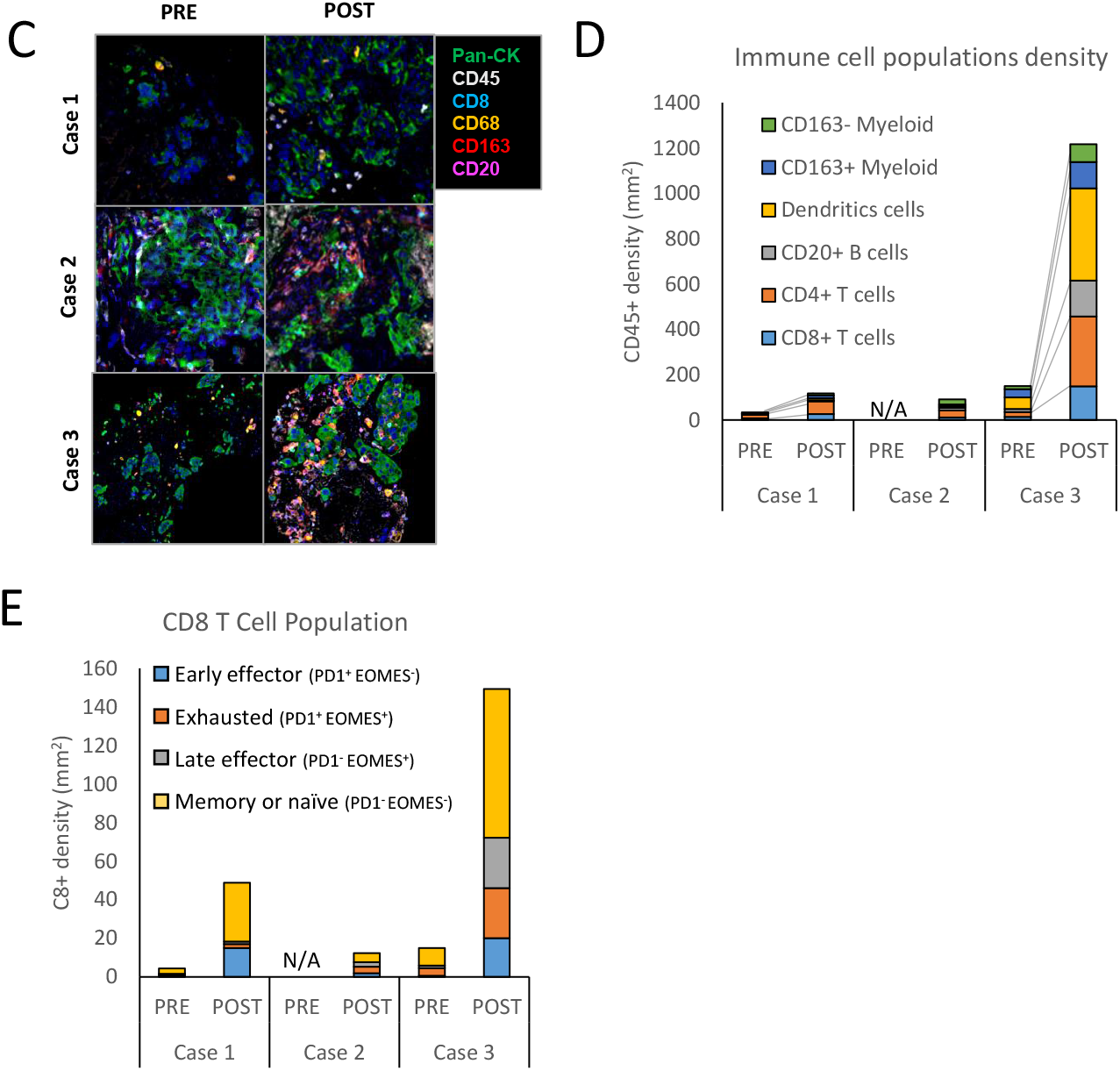
Tumors histopathological phenotype and immune composition. **(A)** An H&E and **(B)** CLIA IHC staining of estrogen receptor (ER), progesterone receptor (PR), androgen receptor (AR) and human epidermal growth factor receptor 2 (HER2) was performed on both pre and on-treatment biopsies for each patient. mIHC was used to **(C)** reconstruct a region of each tumor showing epithelial cells (pan-CK) and various immune cells subtypes as indicated, **(D)** identification of immune cell population subset and density, and **(E)** determination of CD8^+^ T cell population characteristics based on expression of PD-1 and EOMES expression.

### Reverse-phase protein array

Protein samples were analyzed by reverse-phase protein array (RPPA) as previously described (Tibes et al., 2006, Zhang et al., 2009). For scaling purposes, patient data was normalized to TCGA RPPA breast cancer dataset (Akbani et al., 2014). Heat maps were generated for each patient by ordering proteins from most downregulated to most upregulated in the on-treatment sample (ratio of treated to pre-treatment). Heat maps were generated using programs Cluster 3.0 and TreeView. Pathway scores for each patient sample (and each sample in the TCGA cohort) were analyzed as previously described (Akbani et al., 2014, Labrie et al., 2019b). A list of protein and phosphoprotein RPPA predictors used to calculate each pathway score is available in **Suppl. Table 2**.

### Cyclic-Immunofluorescence

Cyclic-Immunofluorescence (Cyc-IF) allows the detection of more than 40 proteins on a single FFPE slide. Multiple sequential rounds of immunofluorescence staining and quenching were performed on each patient sample as previously described (Lin et al., 2016, Lin et al., 2015, Lin et al., 2018) and a detailed method can be found in **Suppl. Method**. Primary antibody details are listed in **Suppl. Table 3**.

## Results

### Patient characteristics

Three participants with mTNBC were enrolled in the pilot study and successfully underwent pre-treatment biopsy, one cycle of olaparib monotherapy, and a repeat on-treatment biopsy before starting the olaparib and durvalumab combination. **Table 1** depicts the baseline characteristics of each patient. Median age of participants was 51.3 years old. All participants had more than three sites of metastatic disease, including visceral disease. All participants had received either prior neoadjuvant or adjuvant chemotherapy; none had prior systemic therapy for metastatic disease.

**Table 1:**
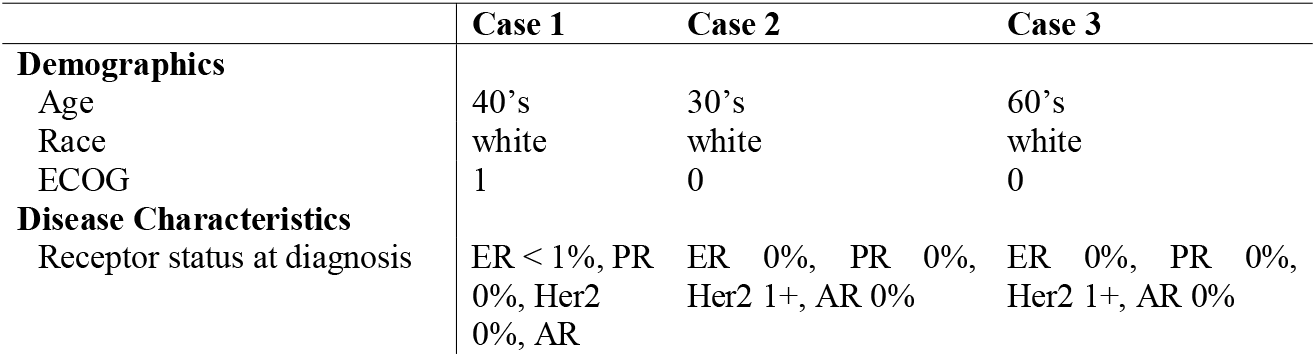
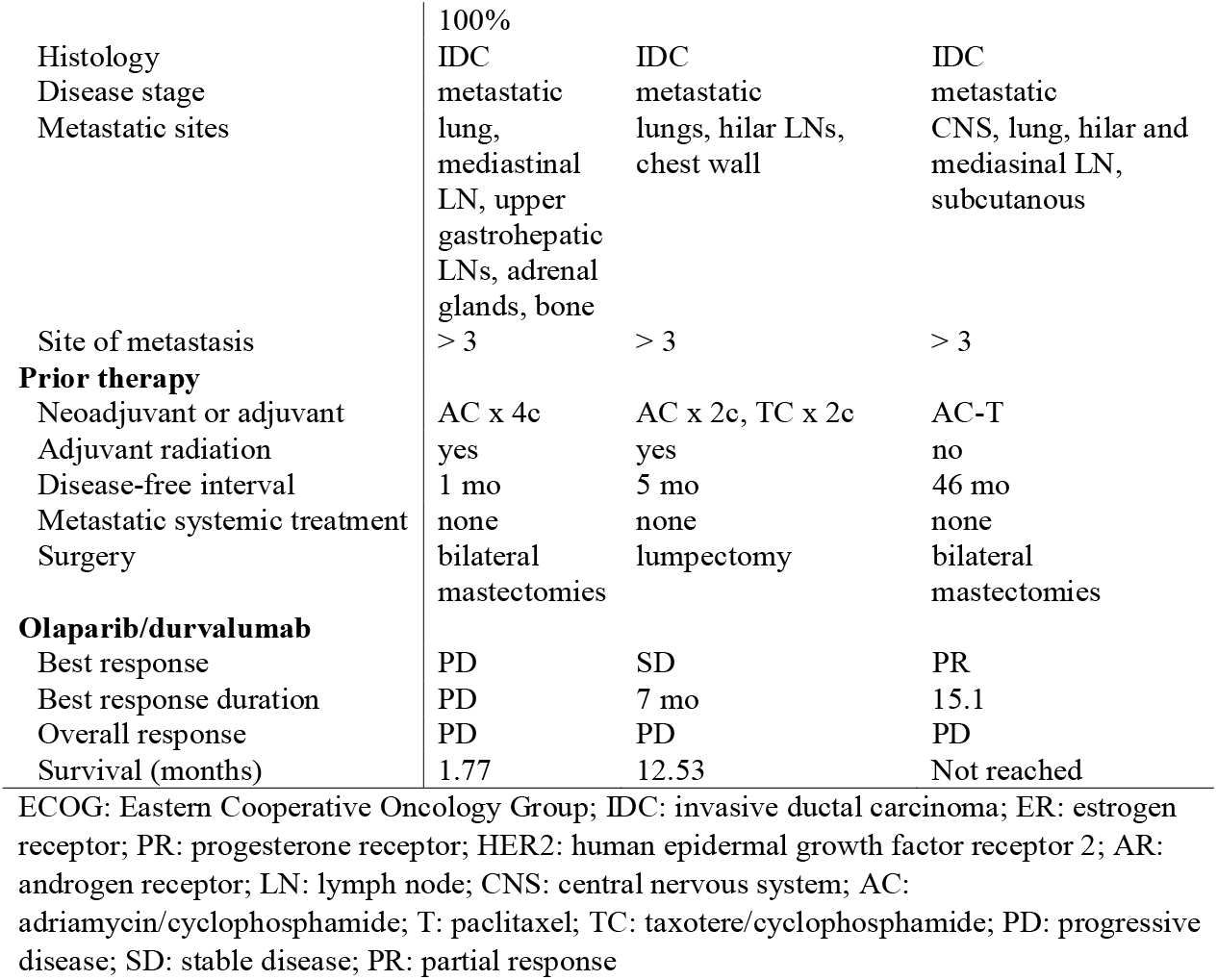
Patient’s characteristics.

### Feasibility

All three participants completed the scheduled pre- and on-treatment biopsies of metastatic tumor. With the exception of two assays, all predetermined CLIA analytics on the pre-treatment biopsy specimens were completed within the previously defined 4-week window prior to the on-treatment initiation biopsy. As shown in **Suppl. Table 4**, the median time of completion was 13 days, meeting the primary endpoint of feasibility of completing CLIA analytics. Given the success for the feasibility endpoint in the first three participants, the study received support to expand to a larger phase 2 efficacy trial. Thus, enrollment on this pilot study was suspended prior to the planned target of 8 participants.

### Safety

There were no adverse events (AE) related to pre- and on-treatment biopsies in all three patients. Only one grade 3-4 event was noted in the patients, which consisted of a hip fracture in a patient with known bone involvement and was considered unrelated to treatment **(Table 2)**. All other events were grade 1-2, with the most common AEs being fatigue (n=2/3), nausea (n=2/3), constipation (n=2/3), and muscular skeletal pain (n=2/3). There were no AEs that led to treatment discontinuation. No immune-related AEs were observed.

**Table 2:** Adverse events.

| **Grade** | **AEs** | **% (n=3)** |
| --- | --- | --- |
| Grade 3-4 | hip fracture* | 33% (1) |
| Grade 1-2 | fatigue | 66% (2) |
|  | neutropenia | 33% (1) |
|  | anemia | 33% (1) |
|  | thrombocytopenia | 33% (1) |
|  | hypocacelmia | 33% (1) |
|  | hypomagnesemia | 33% (1) |
|  | nausea | 66% (2) |
|  | emesis | 33% (1) |
|  | sun sensitivity | 33% (1) |
|  | rash | 66% (2) |
|  | ecchymosis | 33% (1) |
|  | diarrhea | 33% (1) |
|  | constipation | 66% (2) |
|  | muscular skeletal pain | 66% (2) |
|  | headache | 33% (1) |
|  | chills | 33% (1) |

\* Not treatment-related

### Case 1

Caucasian woman in her 40’s with a history of stage I (T1cN0M0) invasive ductal carcinoma (IDC), ER/PR positive, HER2 non-amplified, treated with lumpectomy and SLN evaluation, followed by adjuvant taxotere/cyclophosphamide (TC) chemotherapy for 4 cycles. Upon loco-regional recurrence, a new biopsy showed TNBC disease, and she subsequently underwent bilateral mastectomies and lymph node evaluation, followed by adriamycin/cyclophosphamide (AC) for 4 cycles, and radiation therapy. She presented shortly after radiation therapy with biopsy-proven high grade mTNBC involving lungs, mediastinal and upper gastrohepatic lymph nodes, right adrenal gland, and bones (**Table 1**). The patient received treatment on study, with a best response of progressive disease (PD). The patient developed rapid progression and passed away after 2 months on therapy. During the course of treatment, the patient did not have elevated serum tumor markers (CA15-5, CA27-29, or CEA). Two lesions remained stable, one decreased and one increased in size **(Fig. 1B)**. Both pre- and on-olaparib biopsies were acquired from a mediastinal lymph node. H&E staining of both biopsies showed similar histopathological features, with over 90% tumor content and little immune cell infiltration **(Fig. 2A)**. Pre and on-treatment samples were negative for hormone receptors, except for strong nuclear expression of the androgen receptor (AR). In addition, the on-treatment sample had 5% estrogen expressing cells. Ki67 decreased from 50% in the pre-treatment sample to 30% in the on-treatment sample. PD-L1 was positive low in both samples (1-10% and 10-20% respectively) **(Table 3, Fig. 2B)**. GeneTrails^©^ Comprehensive Tumor Panel and WES showed mutations in *TP53* and *FGFR4* variant of unknown significance (VUS) as well as amplification of *CCND1* and *FGF3* **(Table 3)**. The full list of SNVs and CNVs from WES can be found in **Suppl. Table 5 and 6**, respectively.

**Table 3:**
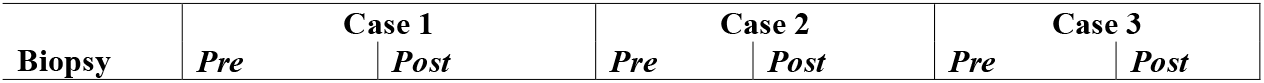
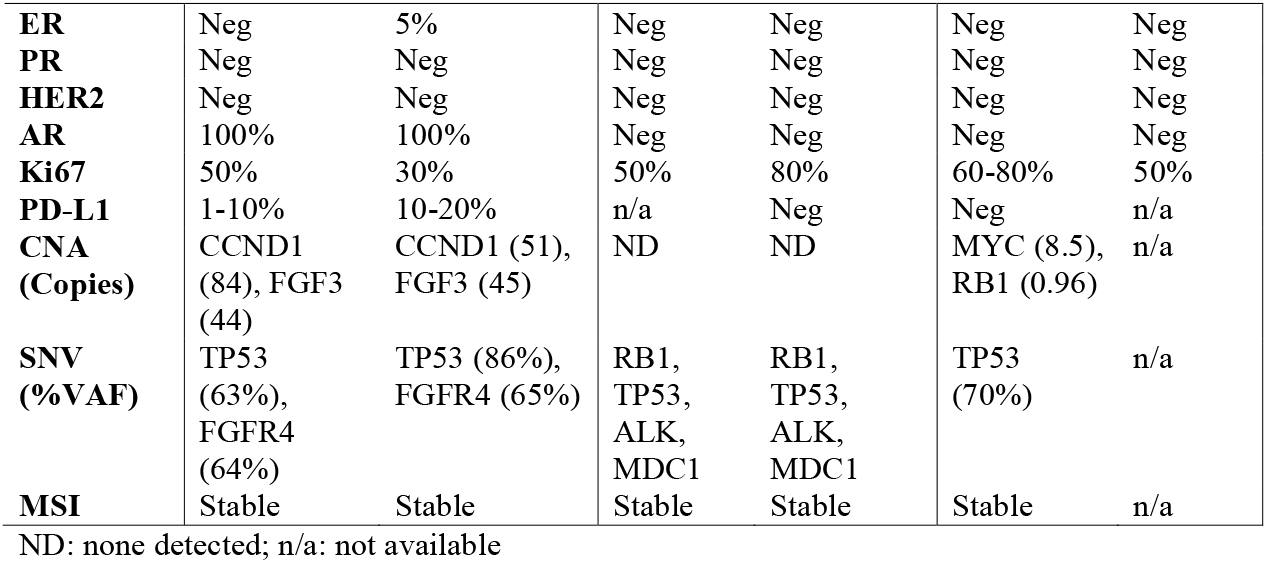
Tumor’s characteristics.

### Case 2

Caucasian woman in her 30’s with a history of stage IIa (T2N0M0) triple negative IDC, treated with lumpectomy, sentinel node biopsy, AC x 2 cycles (stopped due to toxicity), and taxotere/cyclophosphamide (TC) x 2 cycles. She had a first loco-regional recurrence 4 months after initial therapy, treated with modified radical mastectomy followed by 2 cycles of TC and adjuvant radiation therapy. The patient presented with biopsy-proven high-grade mTNBC involving lungs, hilar lymph nodes, and chest wall (**Table 1**). On study, the best response was stable disease (SD) for 7 months prior to progression, and survival of 12.5 months. During the course of treatment, the patient had gradual increases in CA15-5, CA27-29 and CEA that accelerated rapidly prior to death. Two lung lesions slightly increased in size during therapy **(Fig. 1B)**. Both pre and on-olaparib biopsies were acquired from the lung. H&E stained sections showed similar histopathological features in the pre- and on-therapy samples, with over 90% tumor content and minor immune cell infiltration **(Fig. 2A)**. Both biopsies were negative for hormone receptors. Ki67 increased from 50% in the pre-treatment sample to 80% on-treatment. PD-L1 was negative in the on-treatment biopsy (pre-treatment biopsy was insufficient for testing) **(Table 3, Fig. 2B)**. GeneTrails^©^ Comprehensive Tumor Panel and WES analysis revealed mutations in *TP53, RB1, ALK* VUS, and *MDC1* VUS **(Table 3)**. The full list of SNVs and CNVs from WES can be found in **Suppl. Table 5 and 6**, respectively.

### Case3

Caucasian woman in her 60’s with a history of stage I multi-focal invasive IDC, with three lesions triple negative and one weakly ER positive (11%), treated with bilateral nipple sparing mastectomy, adjuvant AC, and taxol chemotherapy. This was followed by anastrazole adjuvant therapy for the weakly ER positive tumor. She developed local regional recurrence in the chest wall after 2 years. Excisional biopsy showed ER negative, PR negative, HER2 normal invasive ductal cancer. She was treated with carbo/gemcitabine followed by radiation therapy. She did well for 5 years when she presented with a subcutaneous mass in the umbilicus. The biopsy again demonstrated TNBC. Imaging showed evidence of metastatic disease involving brain, lung, hilar and mediastinal lymph nodes. (**Table 1**). The patient was enrolled in the study and experienced a partial response (PR; near complete). She had progression of disease in hilar lymph nodes after 15.1 months on study therapy, and progressive mediastinal lymphadenopathy at 19 months. The patient remained alive at time of data cutoff (21 months). During the course of treatment, serum tumor markers were not elevated. All three lesions that were monitored (lung and mediastinum) reduced in size during the course of treatment **(Fig. 1B)**. Both pre- and on-olaparib biopsies were acquired from the left paratracheal mediastinal lymph node. H&E staining showed a major decrease in tumor content (from 90% to 5%) and a marked increase in immune cell infiltration in the on-treatment sample compared to pre-treatment sample **(Fig. 2A)**. The pretreatment sample was negative for PD-L1 (on-treatment was insufficient for testing), and both pre- and on-treatment samples were negative for hormone receptors (ER, PR). Ki67 was positive in 60-80% of cells pre-treatment and > 50% of cells post-treatment **(Table 3, Fig. 2B)**. GeneTrails^©^ Comprehensive Tumor Panel and WES detected a mutation in *TP53*, amplification of *MYC*, and *RB1copy loss* in the pre-treatment sample (insufficient material was available for on-treatment analysis). The patient had a germline deletion of *BRCA1* exons 13-15 that has been previously linked to familial cancer (Seong et al., 2014) however, the impact of this deletion on protein function is currently unknown. The full list of SNVs and CNVs from WES can be found in **Suppl. Table 5 and 6**, respectively.

### Immune monitoring during olaparib monotherapy

Several studies including our own have shown that DNA damage induced by PARPi can trigger an immune response and enhance ICB therapy efficacy (Chabanon et al., 2019, Ding et al., 2018, Huang et al., 2015, Shen et al., 2019). To determine the impact of olaparib treatment on immune cell populations in tumors, pre- and on-treatment samples from patients 1 and 3, and the on-treatment sample from patient 2, were analyzed by mIHC (insufficient material was available from the pre-treatment sample from patient 2). mIHC allows identification and quantification of different immune cell populations on a single FFPE slide. Furthermore, by analyzing the density of immune cells on the tissue, it is possible to determine if the tumors are immune inflamed, defined as high immune infiltrates (> 250 immune cells/mm^2^) (Keren et al., 2018). In patient 1, as shown in **Fig. 2C-E**, the majority of the immune cells in both biopsies were CD4+ T cells. Although a modest increase in density of CD8 T-cells was observed on-treatment, most were memory or naïve T-cells, with only a small fraction of the CD8+ cells (16 cells/mm^2^) classified as effector cells. Consistent with the progressive disease in this patient, olaparib treatment did not significantly alter the immune composition, at least at this time point. In the case of patient 2, the immune cell population analysis revealed a diverse array of immune cells, with a predominance of CD4+ T-cells and CD163-myelomonocytic cells. The CD8+ T-cell population remained small (12 cells/mm^2^), with few effector cells. In contrast, immune cell population analysis from patient 3 revealed important diversity, with CD4+ T-cells and dendritic cells as the predominant populations. Furthermore, the CD8 T-cell population increased (150 cells/mm^2^), and analysis revealed an increase in both early and late effector cell proportions. These results indicate that PARPi activated an immune response in this patient, that is consistent with the subsequent near CR to the olaparib and durvalumab drug combination. Taken together, these results indicate that immune alterations can be detected within one cycle of PARPi treatment through *in situ* mIHC analysis and have the potential to predict benefit to the drug combination.

### Protein network rewiring and adaptive responses

Emerging studies have demonstrated that protein network rewiring can be observed quickly after initiation of PARPi therapy. Adaptive responses due to rewiring of cells that survive PARPi-induced stress PARPi can represent therapeutic opportunities (Fang et al., 2019, Labrie et al., 2019b, Sun et al., 2017). RPPA analysis was used to compare the pre- and on-treatment protein extracts from cases 1 and 3 (**Fig. 3)** (insufficient material was available from case 2). There was a drastic decrease in protein PARylation in the on-treatment samples, indicating target engagement by the drug. In patient 1, a DNA damage response (DDR) was triggered by olaparib, as shown by increased phospho-H2AX, phospho-RPA32, and histone modification (**Fig. 3A**). Interestingly, the drug did not induce a strong overall protein network rewiring in this patient, as shown by the distribution of the protein fold-change (on-treatment/pre-treatment) following PARPi treatment (**Fig. 3B**). Furthermore, pathway analysis revealed few changes in the on-treatment sample compared to the pre-treatment sample (**Fig. 3C**). Of note, when compared to the TCGA breast cancer cohort, pathway analysis revealed a particularly strong DNA damage checkpoint activity as well as a high cell cycle progression score in both pre and on-treatment samples, indicative of a rapidly growing tumor with replication stress (RS). This, combined with the lack of an immune effector infiltrate (Fig. 2) may indicate greater sensitivity to PARP combined with DNA damage checkpoint inhibitors than to the PARP and anti-PDL1 combination the patient received (Fang et al., 2019). Signaling pathways were highly activated in the baseline biopsy from this patient (RTK, RAS-MAPK, PI3K-AKT, TSC-mTOR), with a slight decrease in the on-treatment sample compared to pre-treatment. Immune score correlated with the multiplex IHC showed a low immune cell infiltration in both pre and on-treatment samples. Taken together, these results suggest that while PARPi inhibited its target in patient 1, the tumor remained mostly indifferent to the effects of PARP inhibition.

**Figure 3:**
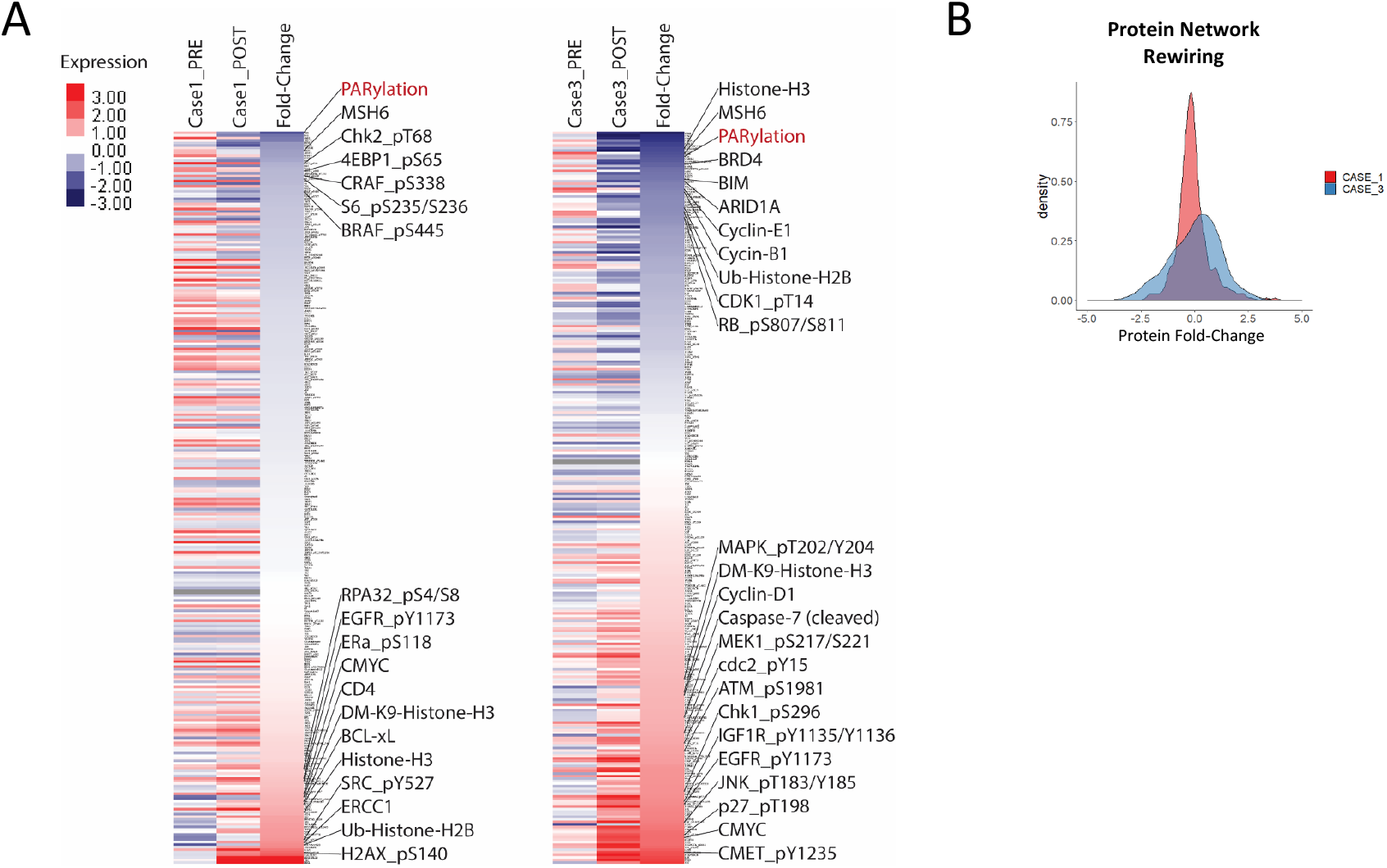
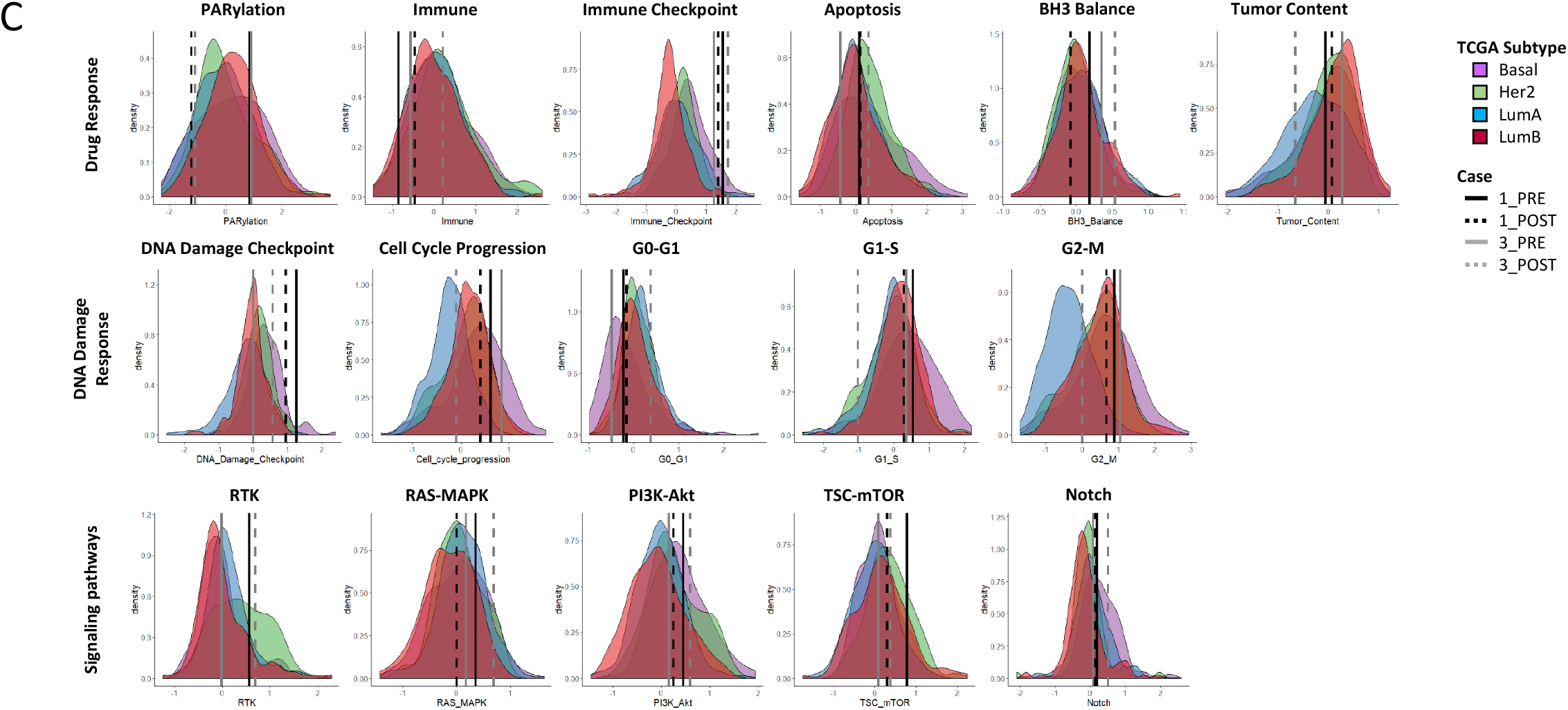
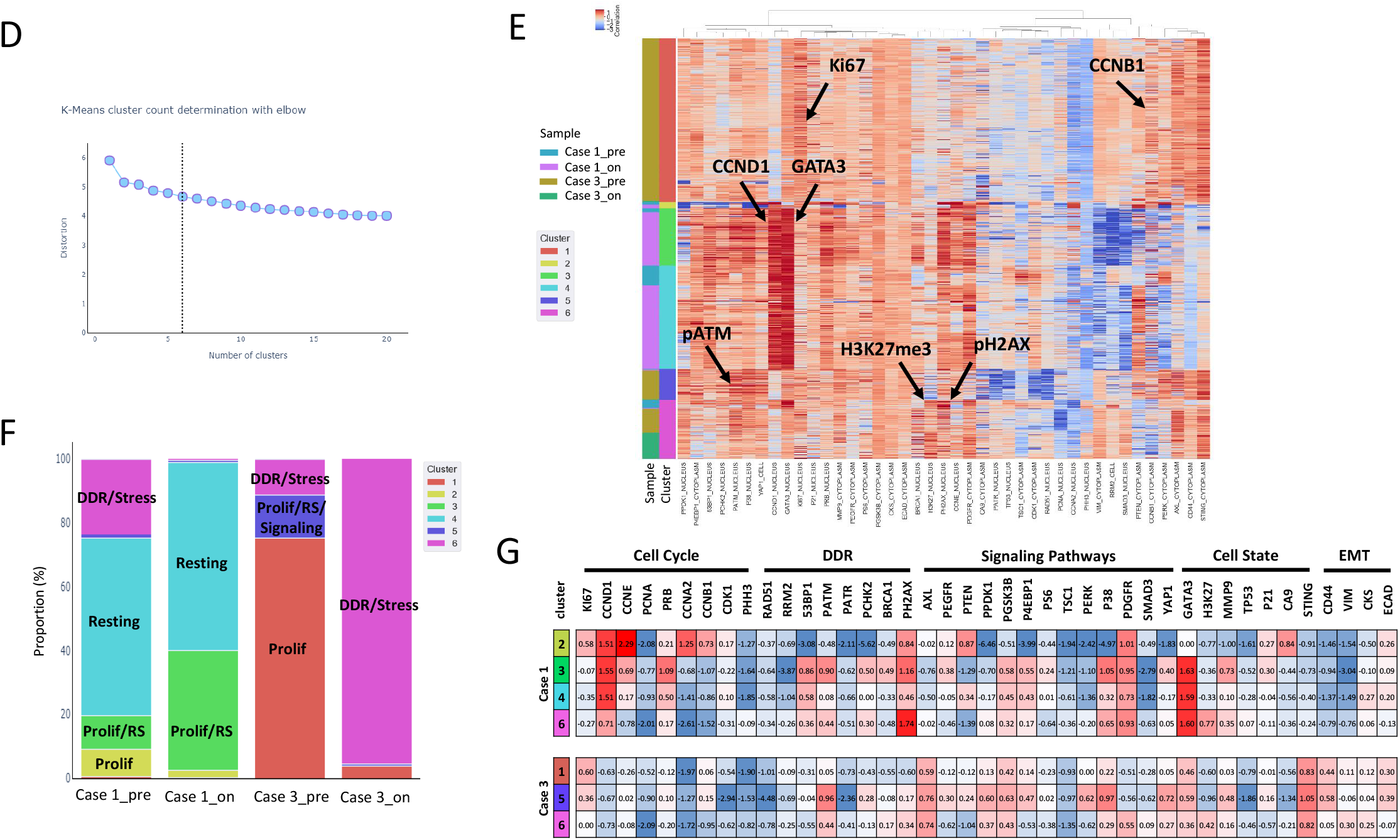
Protein network rewiring and single-cell proteomics analysis of the tumor composition. RPPA analysis was used to analyze protein changes following olaparib treatment. **(A)** Heat map representing the protein expression in each sample as well as the fold change between on and pre-treatment samples from individual patients. The protein was ordered from the most downregulated to the most upregulated in the on-treatment samples compared to pre-treatment. **(B)** Distribution plot showing the fold change of each protein during the course of treatment (on-treatment/pre-treatment). **(C)** Pathway score analysis showing the score distribution of each TCGA breast cancer subtype and the pathway score of each sample from individual patients. Cyc-IF analysis showing different cancer cell populations. **(D)** Chart constructed using the elbow method to determine the optimal number of clusters for **(E)** a K-mean clustering analysis. **(F)** Distribution plot showing the percentage of cells from each K-mean cluster within each sample. **(G)** Table showing the median expression of each protein within each cluster. The table was colored based on expression intensity, with red being high and blue being low expression values.

By contrast, RPPA analysis of patient 3 samples revealed major protein network rewiring, which consisted of alteration of major signaling pathways, immune activity, and cell cycle proteins (**Fig. 3A,B**). Pathway analysis revealed increased immune cell infiltration, DNA damage checkpoint activation, apoptosis, and increased RTK and RAS-MAPK signaling pathways. Together these changes suggest that multiple PARPi combination therapies could lead to patient benefit. In addition, a markedly decreased tumor content and cell cycle progression was observed, consistent with the tumor being responsive to the PARPi (**Fig. 3C**).

Cyc-IF single cell proteomics analysis was used to study changes in tumor heterogeneity and cell state in response to olaparib treatment **(Fig. 3D-G, Suppl. Fig. 1)**. K-means clustering **(Fig. 3D,E)** of pre and on-treatment samples from both patient 1 and 3 was performed, using the elbow method to determine optimal cluster number. As shown in the heat map, patient 1 displayed high levels of cyclin-D1 and GATA3, supporting findings from DNA sequencing and IHC staining, which indicated an amplification of the *CCND1* gene and elevated expression of AR, respectively. Indeed, GATA3, which is a known target of the ER, has recently been associated with AR expression in TNBC tumors (Kim et al., 2016, Naderi and Hughes-Davies, 2008). Each of the 6 identified clusters were enriched for different cell populations: (1-2) proliferative, (3) proliferative with RS, (4) resting, (5) proliferative with RS and high protein signaling activity, and (6) DDR and stress. **Fig. 3F,G** illustrate the frequency of different clusters and median expression of each marker across the clusters.

Following olaparib treatment, patient 1 displayed a slight change in population frequencies, with a decrease in the proliferative (high Ki67 and cyclins) and DDR/stress populations (low Ki67 and high DDR proteins) and an increase in proliferating cells that show signs of RS (high DDR proteins, moderate Ki67, and cyclins). More importantly, the resting population (low Ki67, cyclins and DDR proteins, high epithelial markers) remained unchanged, indicating an indifference of this population of cells to olaparib. In patient 3, a major restructuring of cell population frequencies was observed following olaparib treatment. In the pre-treatment sample, about 75% of tumor cells were part of the proliferating cluster (high Ki67, low DDR proteins). The remaining 25% were either part of the proliferative/RS/signaling cluster (moderate DDR and cyclins, high signaling activity, and high MMP9 expression) or the DDR/stress cluster (low Ki67 and cyclins, high DDR, and high H3K27me3). In the on-treatment sample, over 90% of the remaining tumor cells became part of the DDR/stress cluster, indicating a strong response olaparib. Whether this represents a state change or a selection of particular cell populations remains to be determined.

Taken together, baseline and induced changes in RPPA and Cyc-IF may improve the ability to predict tumor response to olaparib, with protein network and cell population changes indicating sensitivity to the effects of PARP inhibition. Indeed, these assays demonstrated a major tumoral restructuring in patient 3, consistent with the tumor ecosystem being sensitive to the effects of PARP inhibition. Conversely, despite inhibition of PARP, patient 1 displayed little protein alteration, suggesting an indifference of the tumor to the effects of PARP inhibition. Further studies with a larger cohort of patients will be needed to determine whether marked changes in protein and immune networks on PARPi therapy portends marked patient benefit.

## Discussion

The main objective of this study was to establish the feasibility of longitudinal multi-omics analysis of serial tumor samples in real-time. Here we demonstrated that: **(1)** Combination olaparib and durvalumab therapy can be administered safely in mTNBC patients, **(2)** Serial tumor and blood samples can be collected with minimal risk and analyzed within a 28-day window with CLIA assays, **(3)** Multi-omics analysis of these samples results in complementary information that can help characterize basal states and adaptive responses of the tumor ecosystem to olaparib, **(4)** changes in tumor state and immune composition following olaparib treatment can be detected within one olaparib cycle and may predict benefit from therapy, and (**5**) changes in the tumor and ecosystem may aid in the identification of PARPi combinations that could result in therapeutic benefit.

Our previous studies demonstrated that PARPi monotherapy triggers adaptive responses early during the course of treatment, which represents therapeutic opportunities and can be analyzed through the collection of serial samples (Labrie et al., 2019b). This motivated sample collection during PARPi treatment, which would potentially enable us to identify early markers of response and resistance to olaparib. Multi-omics analysis of serially collected samples clearly showed changes in the tumor composition and cell states following therapy initiation confirming our previous observations. Interestingly, there was a strong consensus across the data obtained from different assay types and complementary supporting information that helped illustrate the complexity of each mTNBC tumor and response to treatment. The trial design of a monotherapy run-in of olaparib with an on-treatment biopsy was particularly important in determining the consensus across multi-platform assays as well as the robustness and sensitivity of each assay. For example, immune cell infiltration observed by H&E in the on-treatment biopsy from patient 3 correlated with RPPA data analysis and mIHC. The mIHC assay added an important layer of information by accurately describing the different immune cells populations providing a snapshot of immune contexture. As for the genetic alterations that were observed through DNA sequencing, we could correlate mutations and copy number alterations with the protein expression data, using RPPA and Cyc-IF. Finally, RPPA and Cyc-IF proteomic assays were also shown to be complementary, whereby RPPA helped identify important pathway alterations in each patient sample, while Cyc-IF helped determine the proportion of cancer cells that displayed these features. This data integration will be particularly important in the next phase of our study to develop new CLIA assays to predict response to therapy.

Although mTNBC is a heterogeneous group of diseases with various molecular aberrations, it is usually treated as a single group, likely contributing to the limited efficacy observed across multiple trials (Burstein et al., 2015). Indeed, in this study, the patient in case 1 who had rapid progression was AR positive and likely represented a luminal androgen receptor (LAR) subtype of breast cancer, rather than the basal subtype that dominates TNBC. AR expression in breast cancer has been shown to limit effects of PARPi in preclinical models and may explain the indifference of the tumor in case 1 to PARP inhibition (Li et al., 2017, Min et al., 2018, Luo et al., 2016). Both the MEDIOLA and TOPACIO trials demonstrated activity of PARP and ICB in TNBC. The limited activity particularly in BRCA1/2 wild type tumors may have been influenced by the make-up of the TNBC populations studied. Whether the benefit was limited to patients with basal breast cancer lacking AR expression was not determined in these studies.

PARPi and ICB monotherapies have shown encouraging results in various cancers, but their efficacy is limited in mTNBC patients because of the rapid development of resistance. Previous studies have shown that PARP inhibitors have immunomodulatory properties that have been proposed to increase tumor antigens, antigen presentation, and cytotoxic T-lymphocyte recruitment (Galluzzi et al., 2012, Huang et al., 2015), and upregulate PD-L1 expression through induction of stimulator interferon genes (STING) response and interferon production (Jiao et al., 2017, Chabanon et al., 2019, Ding et al., 2018, Shen et al., 2019), all of which may increase the efficacy of ICB therapy and extend PARPi efficacy beyond tumors with intrinsic homologous recombination DNA repair deficiency (HRD).

In this pilot study, one of the three patients treated with the drug combination had a stable disease for more than six months and another patient had a near complete response that was sustained for almost two years, indicating that a subset of mTNBC patients and more specifically basal breast cancers with both wild type and mutant BRCA genes could benefit from this drug combination. There is an urgent need to better identify patients that are most likely to benefit from this drug combination. Further, for those patients who are not predicted to benefit from the PARPi and ICB combination, alternative therapy approaches are needed. For those patients whose tumors are unresponsive to inhibition of PARPi, it may be optimal to move from PARPi to an alternative drug or drug combination. For those patients for whom there are marked effects of PARPi but no justification for ICB, it may be optimal to add other drugs to the PARPi, such as AKT or MAPK pathway inhibitors, or DNA damage checkpoint inhibitors, based on changes in the tumor microenvironment. Indeed, we have demonstrated benefit of these combinations in a number of model systems as well as in subpopulations of patients (Fang et al., 2019, Konstantinopoulos et al., 2019, Matulonis et al., 2017, Sun et al., 2017). The benefit of the combinations in a subpopulation of models and patients supports the concept that if biomarkers that can direct patients to particular combinations can be identified on the pre- or on-therapy biopsy, patient benefit from receiving the appropriate combination may be greatly increased. In this study, for example, patient 1 demonstrated no benefit from the olaparib and durvalumab combination; however, benefit might have been seen from the combination of PARPi and DNA damage checkpoint inhibition, based on marked replication stress and DDR activation (Fang et al., 2019).

Inherent resistance, characterized by a pre-existing resistance to therapy, is difficult to predict in static biopsies that are acquired prior to therapy. We and other groups have demonstrated in multiple studies that the acquisition of a second biopsy early during the course of treatment is a powerful tool for analyzing how the tumor responds to therapy and for helping to identify mechanisms of adaptive resistance (reviewed in (Labrie et al., 2019a)). We have shown that serial samples can be collected pre and on-therapy with minimum risk to the patient and that these samples can be analyzed in “real time” using CLIA assays. Further, we have demonstrated that the serial analysis of tumors under olaparib stress provides information content that is not available from pre-treatment biopsies. The increase in information content available from serial biopsies parallels our results from a window of opportunity trial in ovarian cancer (Labrie et al., 2019b). Indeed, the pathways activated by PARPi in breast cancer cells, as well as the potential combinations indicated by the serial analysis, are remarkably similar in both diseases. The similarity in responses between this study, in which samples were collected at one month, and the ovarian window of opportunity trial, in which samples were collected 10-14 days after initiation of PARP therapy, indicates that biopsies taken earlier in the course of PARP therapy could be informative and allow initiation of rational combination PARPi therapies earlier in patient management. Interestingly, our preliminary data obtained during this pilot study demonstrated a major network restructuring of the tumor for patient 3, who achieved a partial response on therapy, in contrast to the tumor for patient 1, who had progressive disease. This tumor restructuring was observed in multiple assay types (mIHC, RPPA, and Cyc-IF) and did not rely on specific biomarkers, suggesting that changes in tumor states might represent a biomarker of response and further justifies the use of serially collected samples. Although these assays appear to help predict patient response to therapy and could help with clinical decision-making, it will be important to confirm our findings in the larger phase-2 efficacy trials that we are currently conducting.

Based on this pilot study, one of the main challenges that might be encountered during this novel approach is the availability, accessibility, and quality of tissue samples. Unfortunately, as demonstrated by the lack of sufficient tissue from patient 2 to run all planned assays, the quality and amount of tissue available from each biopsy can pose an analytical challenge. Patient 2 had stable disease that could not be clearly characterized through our assays because one of the biopsies did not yield sufficient material to perform RPPA and Cyc-IF. A pre-treatment sample was also not available for immune monitoring through mIHC. The development of additional assays that do not rely on tumor biopsies may increase the probability of being able to identify a patient’s response to therapy and rationally choose a targeted drug combination. The use of serially collected blood samples could represent an opportunity to characterize ctDNA, circulating tumor cells, vesicles, and proteins, and help determine response to therapy. Several studies have shown that serial liquid biopsies can be useful in determining patient response to therapy and can also help identify mechanisms of acquired drug resistance. However, little is understood about the ability to utilize serial liquid biopsies to identify specific adaptive resistance mechanisms of tumor to therapeutic pressure, which would be essential to more successfully identifying potential targeted drug combinations. For these reasons, further studies are needed to confirm if tumor restructuring can be observed from liquid biopsies and if this information can help predict combination therapies.

In conclusion, preclinical models have identified a number of potential PARPi combinations, some of which have been evaluated in early phase clinical trials (Fang et al., 2018, Fang et al., 2019, Ibrahim et al., 2012, Jiao et al., 2017, Juvekar et al., 2012, Konstantinopoulos et al., 2019, Labrie et al., 2019b, Li et al., 2017, Liu et al., 2013, Matulonis et al., 2017, Min et al., 2018, Shen et al., 2019, Sun et al., 2017, Sun et al., 2018, Vena et al., 2018, Wang et al., 2019, Yin et al., 2017). However, responses, while encouraging in a subset of patients, remain limited in depth and duration. Additionally, a major limitation of prior trials is relying on archival or pre-treatment tissue, which limits the evaluation of dynamic tumor and tumor ecosystem changes in response to therapy. This pilot study demonstrates the feasibility and the importance of using serial samples to analyze tumor response to olaparib monotherapy in real-time to inform on biomarkers to select patient specific olaparib combination therapies. Deep analysis of serial biopsy samples identified biomarkers that could be used to select patient specific combinations of PARPi with ICB, PARPi and AR in LAR patients, PARPi and DNA damage checkpoint inhibitors targeting ATR, CHK1 or WEE1, or PARPi and inhibitors targeting RTK, PI3K-AKT and RAS-MAPK signaling pathways. These observations need to be confirmed in a larger precision oncology prospective trial such as that we have initiated at the Knight Cancer Institute to refine the utility of dynamic biomarkers of response for particular PARPi combinations. Furthermore, the deep analysis of longitudinal biopsies is expected to identify new mechanisms of response and resistance to therapy as well as novel PARPi combinations that can explored in future trials.

## Data Availability

There is no external datasets or online supplementary material.

## Acknowledgments

We gratefully acknowledge the participation, support, and contribution of the SMMART patient advocates Dottie Waddell and Marilyn McWilliams. We also thank the Prospect Creek Foundation and members of the Knight Diagnostic Laboratories (Tanaya Neff, Carol Beadling, Jen Robbins), Knight Biolibrary (Danielle Galipeau) and the Histopathology Shared Resource (Todd Camp).

## Supplementary method

GeneTrails^©^ Comprehensive Solid Tumor Panel, Multiplex Immunohistochemistry, Image Acquisition and Processing, Cyclic-Immunofluorescence, Image processing and analysis for Cyclic-Immunofluorescence.

**Supplementary Table 1: mIHC antibody panel**.

**Supplementary Table 2: RPPA pathway score predictors**.

**Supplementary Table 3: Cyc-IF antibody panel**.

**Supplementary Table 4: Time of completion of the CLIA assays**. The time of completion was measured from the time of sample collection to the time of clinical report completion. N/a: no data available due to low tumor content.

**Supplementary Table 5: List of SNVs detected through WES**.

**Supplementary Table 6: List of CNVs detected through the CNV kit**.

**Supplementary Figure 1: Cyc-IF staining example**.

